# Genomic summary statistics and meta-analysis for set-based gene-environment interaction tests in large-scale sequencing studies

**DOI:** 10.1101/2022.05.08.22274819

**Authors:** Xinyu Wang, Duy T. Pham, Kenneth E. Westerman, Cong Pan, Alisa K. Manning, Han Chen

## Abstract

We propose an efficient method to generate the summary statistics for set-based gene-environment interaction tests, as well as a meta-analysis approach that aggregates the summary statistics across different studies, which can be applied to large biobank-scale sequencing studies with related samples. Simulations showed that meta-analysis is numerically concordant with the equivalent pooled analysis using individual-level data. Moreover, meta-analysis accommodates heterogeneity between studies and enhances power in multi-ethnic studies. We applied the meta-analysis approach to the whole-exome sequencing data from the UK Biobank and successfully identified gene regions associated with waist-hip ratio, as well as those with sex-specific genetic effects.

## Background

Complex diseases are influenced by the synergy of genes and the environment. The study of gene-environment interactions (GEI) may shed light on disease etiology and help identify environmental risk factors that modify the effects of disease-susceptible genes, as well as genetic variants that modify the effects of environmental risk factors for complex diseases (1,2). Following rapid advances in next-generation sequencing (NGS) technologies over the past years, a growing body of research focuses on investigating GEI effects in rare variants (with minor allele frequency (MAF) < 5%), which provides insights into additional disease risk and trait variability undiscovered in common variants from genome-wide association studies (GWAS) (3,4). As in genetic main effect tests, single-variant tests are underpowered in rare variant GEI tests (1,2). A wide variety of set-based GEI tests were developed to improve the statistical power, which are commonly extensions of the burden test and sequence kernel association test (SKAT) (5-7), or a combination of them (8-10). Additionally, the joint tests of both genetic main effects and interaction effects for rare variants were developed to identify variants associated with complex traits, accounting for heterogeneous genetic effects in different environmental exposures (8,10). It is especially useful to perform joint tests when the main effects are small and GEI effects are difficult to detect through interaction only tests (11,12).

Detecting rare variant GEI effects requires extremely large sample sizes given the low power as only a few individuals carried a rare variant while exposed to the environment factor (2,12). Meta-analysis is a well-established way to achieve the scale of sample size needed by rare variant GEI tests. By aggregating summary statistics from different studies, meta-analysis not only increases the sample size, but also avoids sharing individual-level data, which is usually restricted by study policies and confidentiality laws. On the other hand, the file sizes of individual-level genotype data are often extremely large, making it difficult to transfer across platforms with limited disk space and computational resources. The meta-analysis approach can circumvent this problem easily as it does not require sharing individual-level data but rather the genomic summary statistics which are more resource-efficient.

Meta-analysis tools have been developed for genetic main effect tests (13-21). In GWAS, the widely-used inverse variance weighted meta-analysis for common variants usually combines the single-variant effect estimates and their standard errors (21). Meta-analysis for rare variants cannot use the same approach, as models estimating multiple genetic effect sizes with large variances for sparse data can fail to converge, making the algorithm numerically unstable. For this reason, meta-analysis tools for rare variants usually combine score statistics and covariance matrices of individual variants to recover the set-based tests, which are better suited for low-frequency and rare variants. Among these tools, MetaSKAT (13), MASS (17), and Meta-Qtest (19) were developed for unrelated samples. For related samples, RAREMETAL (15-17) can retrieve either the burden test or SKAT, whereas SMMAT (20) can recover burden, SKAT, and a unified test that combines both.

For set-based GEI tests and joint tests, however, very few meta-analysis methods have been developed. Among them, ofGEM (22) introduces filtering statistics based on meta-analysis, but it is only applicable to unrelated samples and no joint tests for genetic main effects and GEI effects were proposed. A recent study proposed extending the rareGE framework (8) to meta-analysis, which can also be conducted for unrelated samples only (23). It is not computationally efficient to generate summary statistics using this method, as for every variant set, a separate statistical model accounting for genetic main effects needs to be fitted. Moreover, methods have been proposed for meta-analysis of joint tests for genetic main effects and GEI effects (24, 25), but they are only applicable to single variant tests. It remains a technical gap in the field to efficiently generate and utilize genomic summary statistics for set-based GEI and joint tests. Recently, we developed a computationally efficient method, Mixed-model Association test for Gene-Environment interactions (MAGEE), for rare variant GEI and joint tests (10). The goal of this study is to develop a general framework for genomic summary statistics and meta-analysis for set-based GEI and joint tests, which are applicable to both unrelated and related samples as well as both quantitative and binary traits. Since MAGEE does not require fitting a separate model adjusting for the genetic main effects in each testing region, the genomic summary statistics for set-based GEI and joint tests can be generated efficiently even for large samples in a whole-genome analysis.

## Results

### *P* value benchmark

The simulated individual-level genotype data included 100,000 samples and 100,000 genetic variants in 1,000 groups (100 variants per group). In the meta-analysis, a great amount of time can be saved when the sample size is large as summary statistic-based calculations do not depend on the sample size. For example, we directly computed the *p* values from individual-level data of 100,000 samples, which took 1,030 s on a single thread for each simulation replicate with 100,000 variants. In contrast, when using the summary statistics, computing the *p* values of the same tests took only 9.7 s using a single thread on the same computing server, which saved over 99% CPU time.

For both quantitative and binary traits with different sample sizes, we compared the *p* values from the meta-analysis (assuming homogeneous genetic effects across studies) with those from pooled individual-level data analysis. Each panel in Figure 1 and Figure 2 displays 10,000 *p* values from quantitative trait analyses with related samples. In the MAGEE framework for individual-level data, we developed two GEI tests, interaction variance component (IV) test and interaction hybrid test using Fisher’s method (IF), along with three joint tests, joint variance component (JV) test, joint hybrid test using Fisher’s method (JF), and joint hybrid test using double Fisher’s procedures (JD) (10). We conducted extensive simulations in MAGEE (10), which showed that each test was well-calibrated for type I error rates. As illustrating examples, here we compared results from IF test as the GEI test, and JD test as the joint test, for meta-analysis and MAGEE pooled analysis. The meta-analysis assuming homogeneous genetic effects shared highly consistent empirical GEI and joint test *p* values with MAGEE pooled analysis (Figures 1 and 2), regardless of the simulation scenario of homogeneous or heterogeneous genetic effects, and the precision of estimates increased with the sample size. Most importantly, for small *p* values around the commonly used significance thresholds (5.0×10^−8^ to 2.5×10^−6^), meta-analysis did not lose any accuracy compared with the MAGEE pooled analysis while being more resource-efficient in saving the computational time. We also found similar results for binary traits (Supplemental Figure S1 and S2).

**Figure 1.**
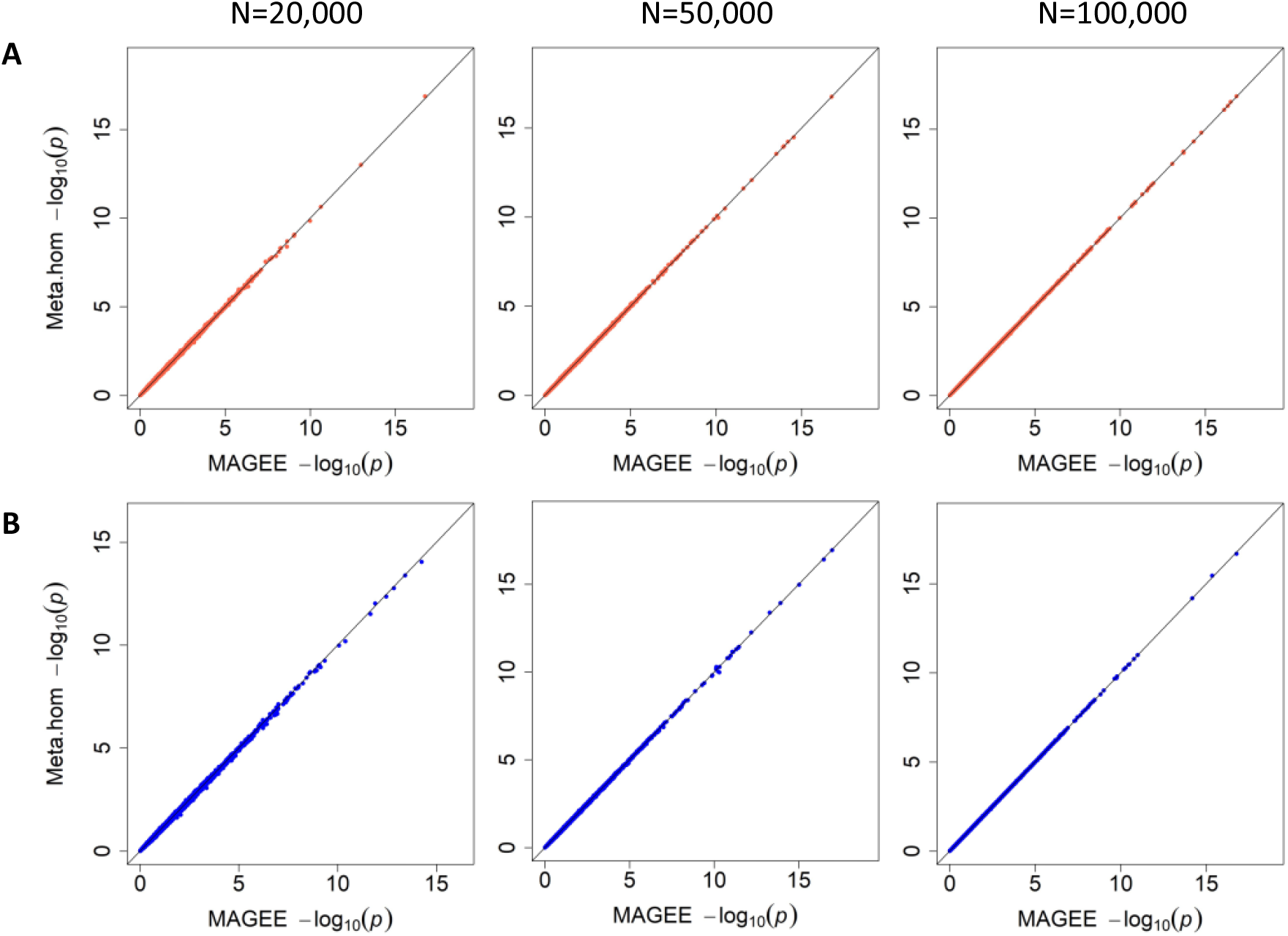
GEI test *p* value benchmark for meta-analysis assuming homogeneous genetic effects with MAGEE in 20,000, 50,000, and 100,000 unrelated samples. (A) Scenario 1 (homogeneous scenario) in quantitative traits. (B) Scenario 2 (heterogeneous scenario) in quantitative traits.

**Figure 2.**
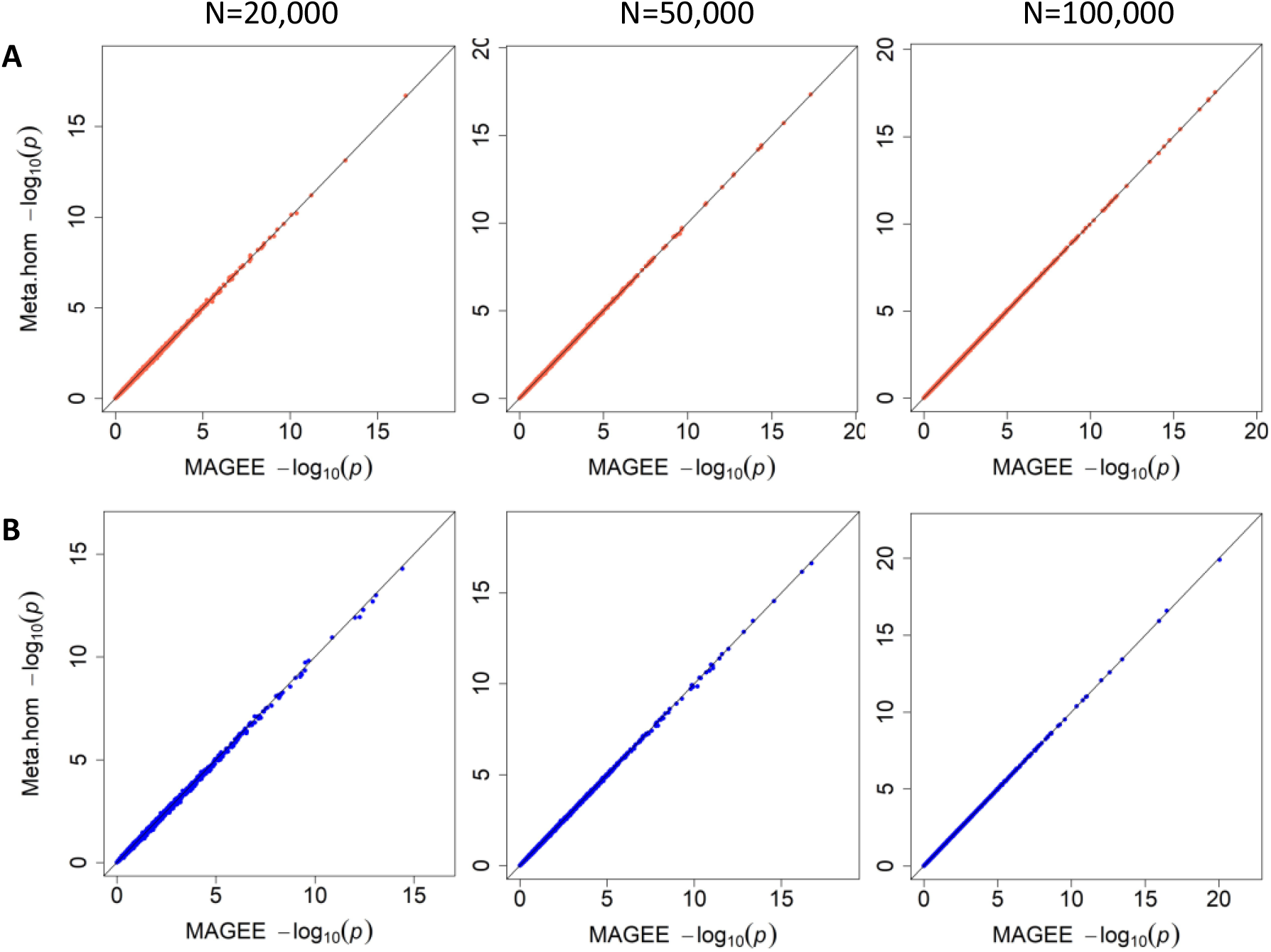
Joint test *p* value benchmark for meta-analysis assuming homogeneous genetic effects with MAGEE in 20,000, 50,000, and 100,000 unrelated samples. (A) Scenario 1 (homogeneous scenario) in quantitative traits. (B) Scenario 2 (heterogeneous scenario) in quantitative traits.

### Power comparison

Figure 3 shows the empirical power of the meta-analysis GEI and joint tests for analyzing quantitative traits from scenario 1, where the two studies had homogeneous covariate effects, genetic effects, and GEI effects. The power was calculated at the significance level of 2.5×10^−6^ with 20,000, 50,000, and 100,000 related samples, respectively. The top panel displays results when the ratio of positive and negative causal variants was 1:1, while the bottom panel displays results when the ratio of positive and negative causal variants was a 4:1. Overall, the meta-analysis tests assuming homogeneous effects are more powerful for both GEI and joint tests, with the IF test being more powerful than the IV test and the JF test being slightly more powerful than the other 2 joint tests.

**Figure 3.**
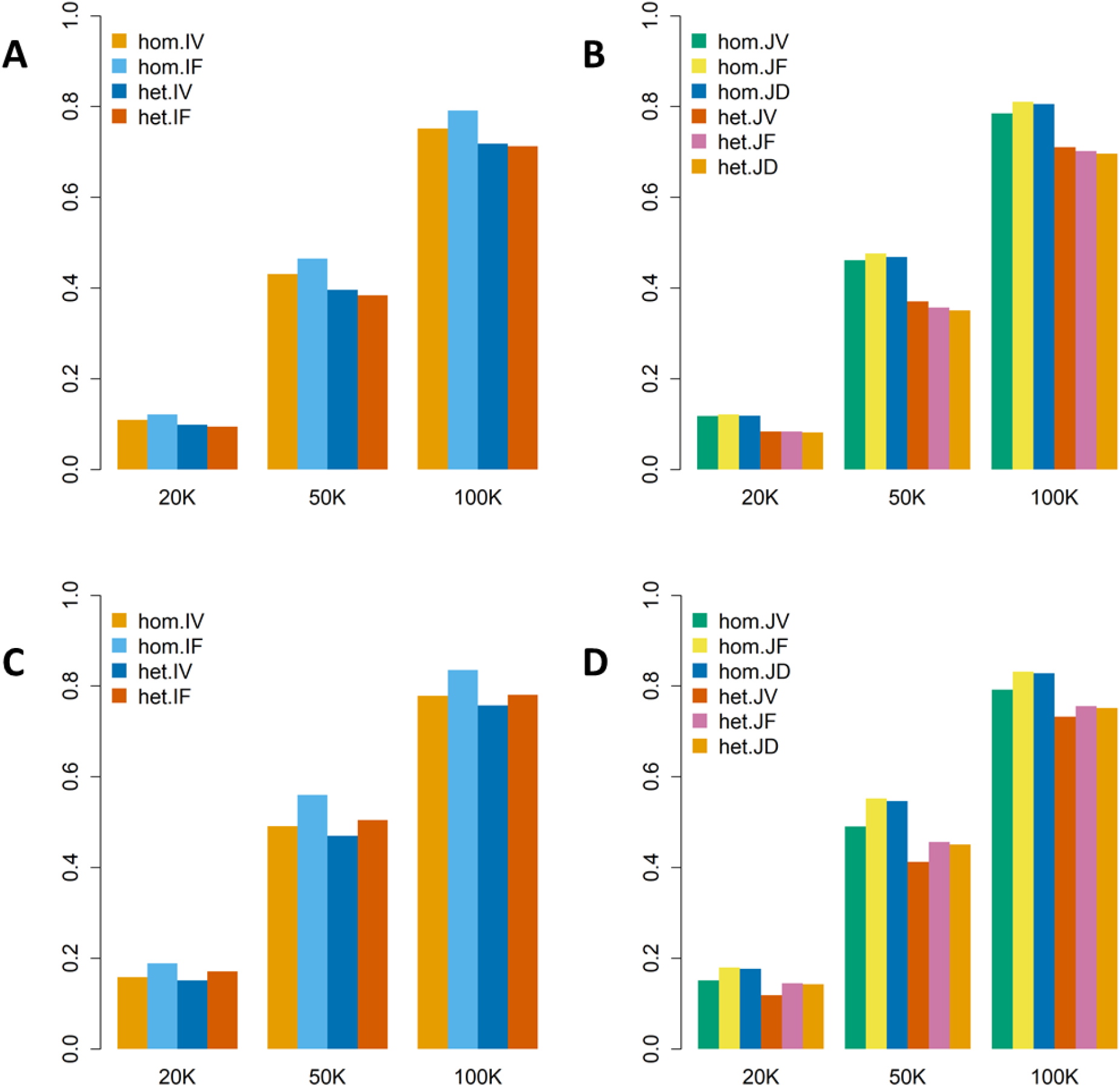
Empirical power of meta-analysis tests for scenario 1 (homogeneous scenario) on quantitative traits in 20,000, 50,000, and 100,000 related samples. (A) GEI tests with 80% null variants, 10% causal variants with positive effects, and 10% causal variants with negative effects. (B) Joint tests with 80% null variants, 10% causal variants with positive effects, and 10% causal variants with negative effects. (C) GEI tests with 80% null variants, 16% causal variants with positive effects, and 4% causal variants with negative effects. (D) Joint tests with 80% null variants, 16% causal variants with positive effects, and 4% causal variants with negative effects.

Figure 4 illustrated the empirical power of the meta-analysis GEI and joint tests for analyzing quantitative traits from scenario 2, where the two studies had heterogeneous covariates effects, genetic effects, and GEI effects. Same as in Figure 3, the ratio of positive to negative causal variants in Figure 4 was 1:1 on the top panel and 4:1 on the bottom panel. In this scenario, as heterogeneity was introduced to these studies, the overall performance of the meta-analysis tests assuming heterogeneous effects outperformed the meta-analysis assuming homogeneous effects for both GEI and joint tests.

**Figure 4.**
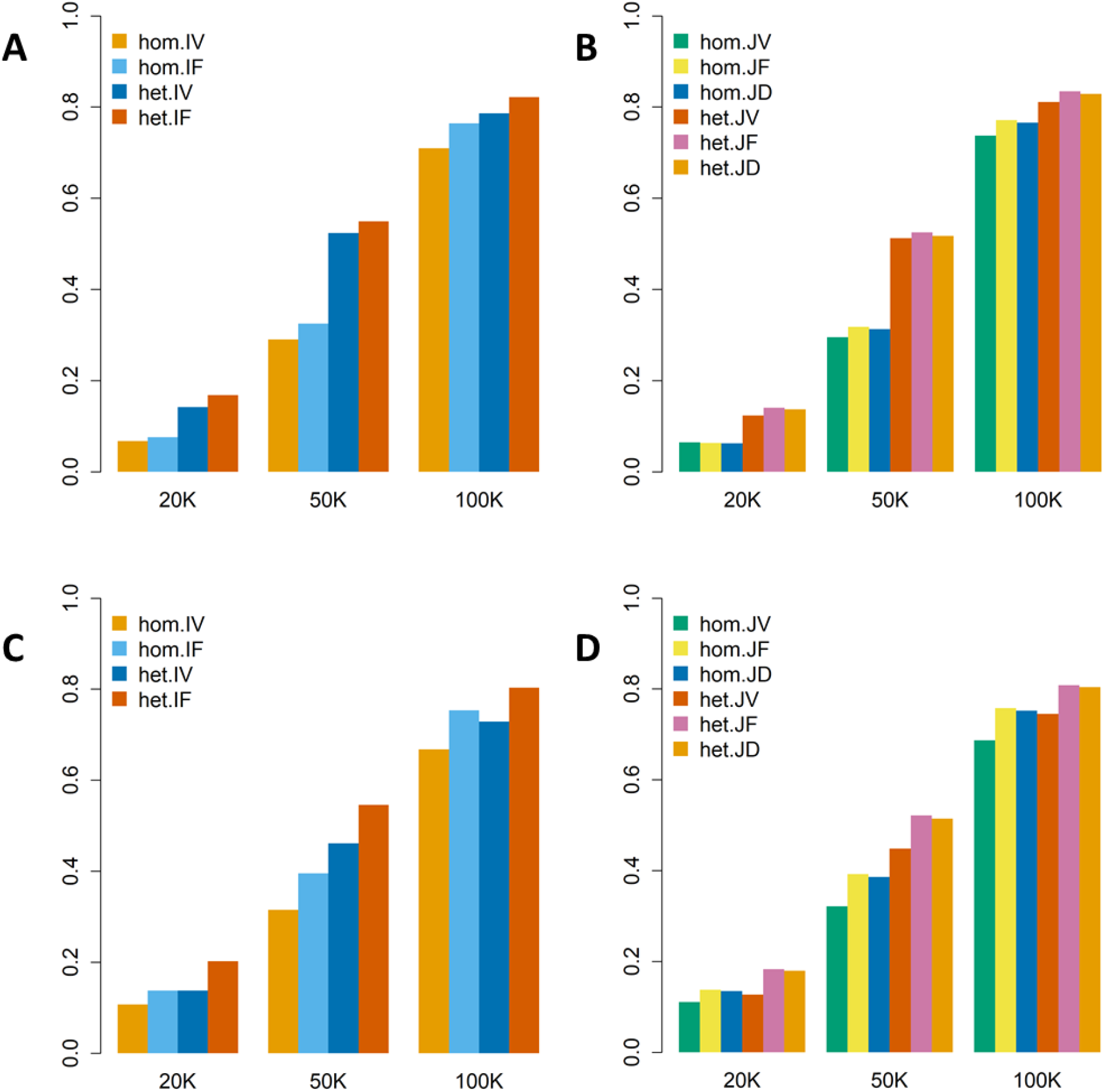
Empirical power of meta-analysis tests for scenario 2 (heterogeneous scenario) on quantitative traits in 20,000, 50,000, and 100,000 related samples. (A) GEI tests with 80% null variants, 10% causal variants with positive effects, and 10% causal variants with negative effects. (B) Joint tests with 80% null variants, 10% causal variants with positive effects, and 10% causal variants with negative effects. (C) GEI tests with 80% null variants, 16% causal variants with positive effects, and 4% causal variants with negative effects. (D) Joint tests with 80% null variants, 16% causal variants with positive effects, and 4% causal variants with negative effects.

The same simulations were conducted for binary traits, and we summarized those results in Supplemental Figures S3 and S4.

### Meta-analysis of waist-hip ratio in the UK Biobank

The original individual-level genotype data from whole-exome sequencing (WES) provided by the UK Biobank for 200,632 samples was 830 GB for 22 autosomes in the PLINK BED format, while the gene-based summary statistics were about 11 GB in total, including 5.4 GB intermediate files from the first batch and 5.4 GB intermediate files from the second batch. By sharing only the summary statistics files for meta-analysis, we saved about 98.7% disk space compared to directly sharing individual-level data for set-based GEI tests. In addition, when we directly computed *p* values from individual-level data, it took 25.48 h (91,732 s) CPU time with 10 threads to conduct all the GEI tests and joint tests in the MAGEE framework. In contrast, we spent only 23.8 min (1,428 s) CPU time using a single thread on the same computing server to compute the *p* values of the same GEI and joint tests when we had the summary statistics, saving about 98.4% of computational time. One additional advantage for meta-analyzing the UK Biobank WES data is that when new samples are released in future tranches, we do not have to spend extra computational cost in rerunning the pooled analysis including previously released samples. Instead, by combining the summary statistics from different releases in a meta-analysis, we can get similar results as the pooled analysis while saving the computational resources.

The two batches of WES data from the UK Biobank were meta-analyzed in 18,668 protein-coding regions to investigate gene-sex interactions effects on waist-hip ratio (WHR). The significance level was adjusted by Bonferroni correction for multiple testing (26) at 0.05/18,668 = 2.7×10^−6^. As shown in Figure 5, the GEI test found three significant regions: *COBLL1* (*p* value = 5.8×10^−10^) on chromosome 2, *HMGA1* (*p* value = 2.8×10^−7^) on chromosome 6, and *VEGFB* (*p* value = 1.3×10^−6^) on chromosome 11. However, none of them remained significant after conditioning on significant variants in the region, defined as those with a single variant joint test *p* value < 5.0×10^−8^, MAF > 1% and genotype missing rate < 5% (Table 1). A total of 13 significant protein-coding regions were found from the joint tests, including *TBX15, ACVR1C, COBLL1, NISCH, PLXND1, CYTL1, HLA-B, HMGA1, KIAA0408, MLXIPL, VEGFB, SBNO1*, and *PAM16*, after excluding less significant protein-coding regions in 1 million base pair flanking regions. Of note, *COBLL1* (*p* value = 1.3×10^−8^ after adjusting for 7 significant single variants) and *KIAA0408* (*p* value = 9.3×10^−9^ after adjusting for 1 significant single variant) had significant joint test *p* values in the conditional analysis (Table 1).

**Table 1.** Meta-analysis results for gene-sex interaction effects on WHR from the UK Biobank WES data analysis.

| Gene | Chr | Start | End | N variants | N sig variants* | Unconditional GEI | Unconditional Joint | Conditional GEI | Conditional Joint |
| --- | --- | --- | --- | --- | --- | --- | --- | --- | --- |
| *TBX15* | 1 | 118883046 | 118989556 | 82 | 1 | 0.79 | **$5.5 \times 10^{-11}$** | 0.67 | 0.011 |
| *ACVR1C* | 2 | 157526767 | 157628864 | 104 | 0 | 0.0089 | **$3.1 \times 10^{-8}$** | 0.0089 | **$3.1 \times 10^{-8}$** |
| *COBLL1* | 2 | 164653624 | 164843679 | 221 | 7 | **$5.8 \times 10^{-10}$** | **$4.2 \times 10^{-16}$** | $3.8 \times 10^{-5}$ | **$7.4 \times 10^{-8}$** |
| *NISCH* | 3 | 52455118 | 52493068 | 348 | 11 | $5.6 \times 10^{-4}$ | **$7.3 \times 10^{-11}$** | $5.0 \times 10^{-4}$ | $3.6 \times 10^{-4}$ |
| *PLXND1* | 3 | 129555175 | 129606818 | 423 | 14 | $4.6 \times 10^{-5}$ | **$3.0 \times 10^{-15}$** | 0.037 | $8.3 \times 10^{-4}$ |
| *CYTL1* | 4 | 5014586 | 5019458 | 33 | 0 | 0.085 | **$1.6 \times 10^{-6}$** | 0.085 | **$1.6 \times 10^{-6}$** |
| *HLA-B* | 6 | 31269491 | 31357188 | 639 | 3 | 0.41 | **$7.5 \times 10^{-12}$** | 0.45 | $2.0 \times 10^{-5}$ |
| *HMGA1* | 6 | 34236873 | 34246231 | 67 | 3 | **$2.8 \times 10^{-7}$** | **$7.3 \times 10^{-28}$** | 0.42 | 0.10 |
| *KIAA0408* | 6 | 127438406 | 127459389 | 76 | 1 | $4.5 \times 10^{-6}$ | **$4.4 \times 10^{-17}$** | 0.0036 | **$9.3 \times 10^{-9}$** |
| *MLXIPL* | 7 | 73593194 | 73624543 | 202 | 4 | 0.0014 | **$5.6 \times 10^{-8}$** | 0.016 | 0.0022 |
| *VEGFB* | 11 | 64234538 | 64238793 | 81 | 2 | **$1.3 \times 10^{-6}$** | **$1.2 \times 10^{-17}$** | 0.22 | 0.16 |
| *SBNO1* | 12 | 123289109 | 123364847 | 344 | 0 | 0.021 | **$1.3 \times 10^{-8}$** | 0.021 | **$1.3 \times 10^{-8}$** |
| *PAM16* | 16 | 4331549 | 4355607 | 234 | 0 | 0.11 | **$1.1 \times 10^{-6}$** | 0.11 | **$1.1 \times 10^{-6}$** |

\* Number of significant variants in the region, defined as those with a single variant joint test $p$ value $< 5.0 \times 10^{-8}$ , MAF $> 1\%$ and genotype missing rate $< 5\%$ . Conditional analysis $p$ values were computed by adjusting for both the genetic main effect and gene-sex interaction terms of significant single variants.

**Figure 5.**
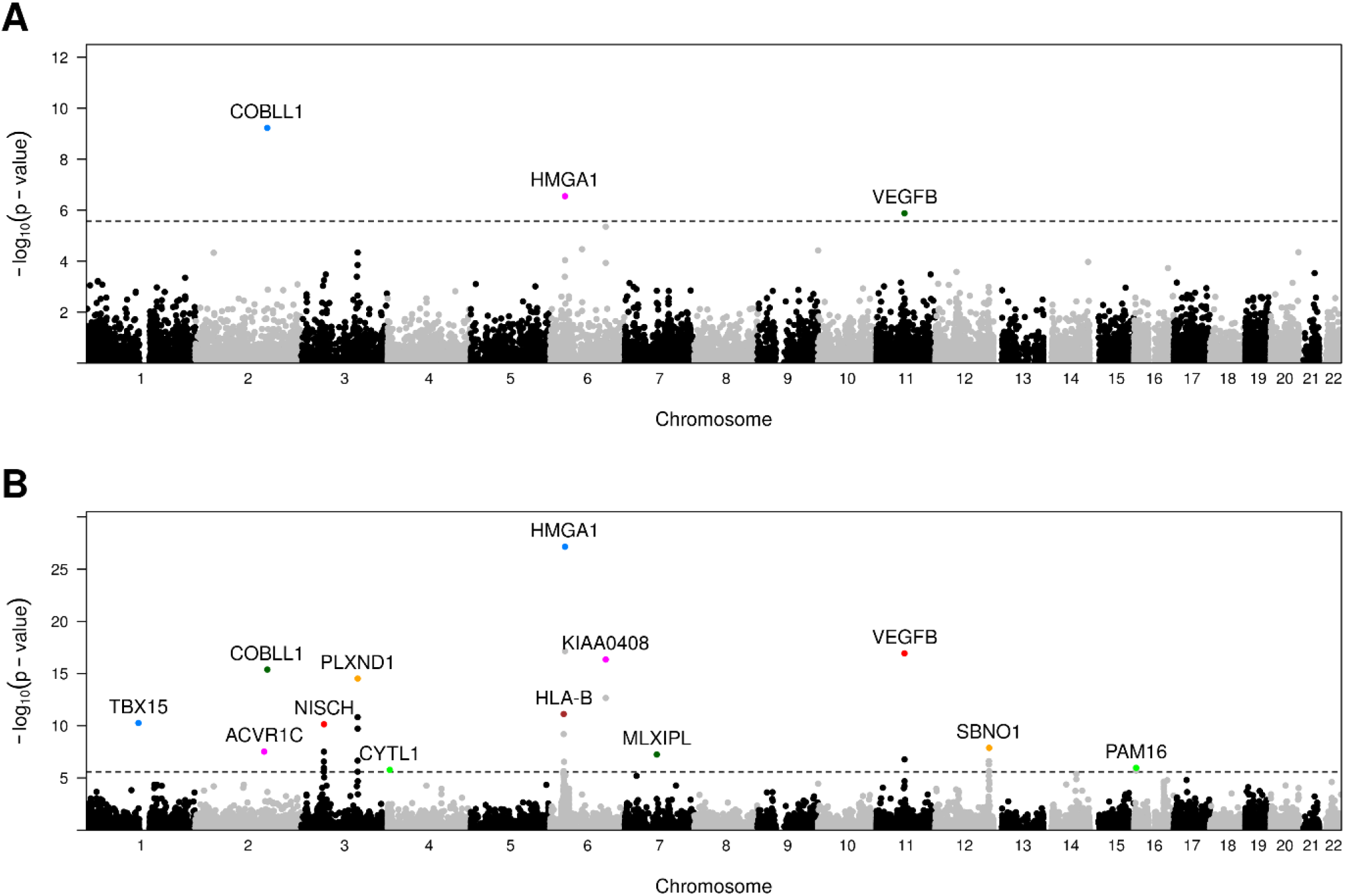
Manhattan plots of UK Biobank WES data analysis using meta-analysis for gene-sex interaction effects on WHR. (A) GEI test. (B) Joint test.

Previous GWAS have identified many loci in these genes for WHR, some of which have sex-dimorphic effects. For example, there is some evidence that certain loci at *COBLL1, PLXND1, HMGA1*, and *VEGFB* have more impact on BMI-adjusted WHR among women (27, 28). In our meta-analysis GEI tests, *PLXND1* did not show significance, however, we did identify gene-sex interactions in the other three regions. Additionally, Justice *et al*. (29) reported gene-sex interactions in *COBLL1, PLXND1*, and *KIAA0408*. There were no significant gene-sex interaction signals in *PLXND1* (*p* value = 4.6×10^−5^) or *KIAA0408* (*p* value = 4.5×10^−6^), possibly because of the limited sample size in our analysis, and more weights were added to rare variants using the beta function, which had potentially diminished the signals from common variants.

We observed most signals driving our joint analyses were contributed by genetic main effects, as only *COBLL1, HMGA1* and *VEGFB* were significant in the GEI tests (they were significant in the main effect tests as well). Several loci in the genes identified in the joint tests were reported to be associated with WHR in previous GWAS analysis (27-32) except for *HLA-B. HLA-B* is a member of the human leukocyte antigen (*HLA*) complex gene family, which helps the immune system distinguish between proteins created by the body and proteins made by outside invaders like viruses and bacteria (33). Further research is needed to investigate if there are any sex-specific genetic impacts on WHR in *HLA-B*. The underlying mechanisms for most of these genetic and sex differences have yet to be discovered.

We pooled individual-level data and ran MAGEE GEI and joint tests to validate the findings. Supplemental Figure S5 shows that *p* values from the meta-analysis and the MAGEE pooled analysis were highly concordant.

Number of significant variants in the region, defined as those with a single variant joint test *p* value < 5.0×10^−8^, MAF > 1% and genotype missing rate < 5%. Conditional analysis *p* values were computed by adjusting for both the genetic main effect and gene-sex interaction terms of significant single variants.

## Discussion

We present a framework for meta-analysis of rare variant GEI and joint tests for continuous and binary traits. The meta-analysis is based on a generalized linear mixed effect model (GLMM), which can account for relatedness in the samples. Single-variant scores and covariance matrices of individual studies can be efficiently generated from MAGEE. The summary statistics for the meta-analysis can be easily aggregated, which allows combining the evidence from millions of samples collected from multiple large-scale sequencing studies to improve statistical power. Additionally, the proposed meta-analysis framework provides flexible set-based joint tests for genetic main effects and GEI effects, including a SKAT-type variance component test, and two unified tests combining burden and SKAT-type tests. In population-based studies, heterogeneity is possible because different ethnic groups might have different MAFs or effect sizes for the same variants, as well as different levels of environmental exposure. Different ways of collecting and measuring data, genotyping methods, quality-control criterion, and imputation methods make studies heterogeneous (34). The meta-analysis can easily account for the heterogeneity across studies, which is difficult in pooled analyses using individual-level data.

We evaluated the performance of the proposed GEI and joint tests through computer simulations. We first showed that meta-analysis assuming homogeneous genetic effects is numerically concordant with the equivalent pooled analysis using individual-level data. Additionally, the proposed methodology can account for heterogeneity across studies in terms of both covariate and genetic effects. GEI and joint tests assuming heterogeneous genetic effects were generally more powerful than homogeneous-type meta-analyses in the presence of simulated heterogeneity. Overall, IF tests were more powerful than IV tests, and JF tests were the most efficient of the three joint tests, in accordance with what we determined from the single-study numerical simulation of MAGEE.

We assessed gene-sex interaction effects impacting WHR, using GEI and joint tests within the proposed meta-analysis framework on WES data from the UK Biobank. GEI tests identified *COBLL1, HMGA1* and *VEGFB* that are already known to have loci with sex-dimorphic effects on WHR. The meta-analysis joint tests identified 13 protein-coding regions, of which 10 had been previously linked to WHR in GWAS (27-32), including *COBLL1, HMGA1, VEGFB, PLXND1* and *KIAA0408* that were reported to have sex-specific genetic loci (27-29). The proposed meta-analysis GEI test did not reach significance for *PLXND1* and *KIAA0408*, possibly due to the limited sample size. To identify rare variants that have effects modified by the environment, extremely large sample sizes are required. Furthermore, we only examined genetic variants in the exome, but noncoding regions may play a substantial role and may have integrated effects with protein-coding genes in complex diseases and traits (27, 35). Once more WES data from the UK Biobank and more whole-genome sequencing (WGS) data become available from other large-scale consortia in the future, we expect that the meta-analysis will be used to identify more novel genetic pathways contributing to a wide range of complex diseases.

Our summary statistics and meta-analysis framework have a few limitations. All participating studies in the proposed meta-analysis must use a unified group definition file to ensure that the summary statistics are generated consistently across studies. Although all component variants need not be present for all studies, if different studies use different gene-level tests or group definition files, the summary statistics would be incompatible (3). In addition, in certain case-control studies, case/control ratios could be extremely unbalanced, leading to strong heterogeneity across studies (13). Future research could explore these extremely heterogeneous cases in further depth.

## Conclusions

We developed efficient and powerful meta-analysis approaches to overcome the challenges associated with set-based GEI and joint tests for rare variants while allowing for sample relatedness. The meta-analysis leverages study-specific genomic summary statistics, eliminating the need to share individual-level data, and is applicable to both quantitative and binary traits. In addition, the meta-analysis can accommodate heterogeneity among studies. These features allow the analysis of GEI effects across multiple large-scale sequencing studies and can improve the power in identification of novel risk factors for complex traits.

## Methods

### Modeling summary statistics

The meta-analysis of rare-variant GEI and joint tests involves two major steps. In the first step, we perform a single-study analysis to calculate the summary statistics of the single-variant score vector and covariance matrix for each variant. The second step consists of combining the summary statistics and constructing the test statistic for the meta-analysis. The first step can be performed by using the MAGEE software package. Below is a brief illustration of how to generate the summary statistics.

Suppose we have *m* = 1, 2, ⋯, *M* studies, first we need to generate the summary statistics for each individual study. The full generalized linear mixed model (GLMM) for a single study *m* is in the following form:

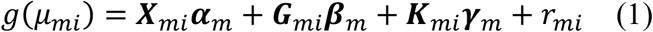

where *g*(·) is the link function of *μ*_*mi*_, which is the conditional mean of the phenotype of individual *i* in study *m*. Typically, for a quantitative trait, *g*(·) is an identity function, while for a binary trait, *g*(·) is a logit function. ***X***_*mi*_ is a vector of *p* covariates including the intercept, and ***G***_*mi*_ is a vector of *q* variants, and ***K***_*mi*_ is a vector of *dq* pairwise GEI terms for *d* environmental factors and *q* variants. Note that the environmental factors are a subset of the covariates in ***X***_*mi*_. Accordingly, ***α***_*m*_ is a *p* × 1 vector for the covariate effects, ***β***_*m*_ is a *q* × 1 vector for the genetic main effects, and ***γ***_*m*_ is the *dq* × 1 vector for GEI effects. The vector for the random intercept 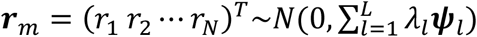, where *N* is the sample size for study *m, λ*_*l*_ are the variance component parameters of *L* random effects, and ***ψ***_*l*_ are *N* × *N* known relatedness matrices.

Through MAGEE, we made statistical inference that the GEI effects test statistic in model (1) can be approximated by fitting a global null model without adjusting for the genetic main effects. Specifically, the global null models are fitted to each individual study as follows:

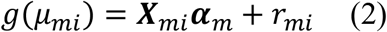

The score vector 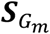 for genetic main effects and GEI effects 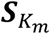 are in the forms of 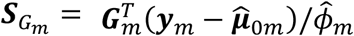 and GEI effects 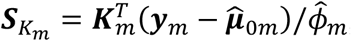 where 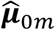 is a vector of fitted values and 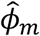 is the dispersion parameter estimated from model (2) in study *m*. Assuming the main effect of genetic variants ***β***_*m*_ are small in a null model with genetic main effects, but without GEI effects *g*(*μ*_*mi*_) = ***X***_*mi*_***α***_*m*_ + ***G***_*mi*_***β***_*m*_ + *r*_*mi*_ (3), we can approximate the score vector for GEI effects accounting for the genetic main effects by 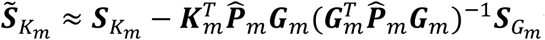 where 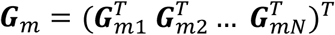 is a *N* × *q* matrix of genetic variants, 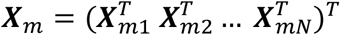is a *N* × *p* matrix of covariates, 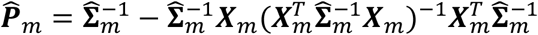 is an *N* × *N* projection matrix from the global null model, where 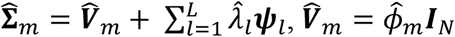 for quantitative traits and *diag*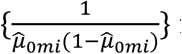 for binary traits, which we estimate from model (2).

In the meta-analysis context, we save the single-variant scores of genetic main effects and GEI effects, 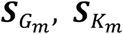, as well as the covariance matrix for the score vectors 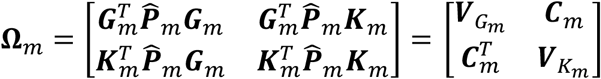 for each participant study *m*, where 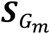 is a *q* × 1 vector, 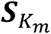 is a *dq* × 1 vector, 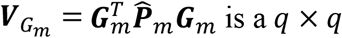 matrix, 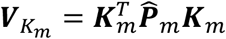 is a *dq* × *dq* matrix, and 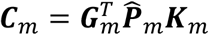 is a *q* × *dq* matrix. As **Ω**_*m*_ is a large and symmetric matrix, we can save the lower triangle elements in a binary format data to save space. Compared to saving the summary statistics as the double-precision floating numbers that each takes 8 bytes, we used 4 bytes to save each floating-point number to further save the space by 50%, without losing accuracy in the downstream meta-analysis.

The test statistics are recovered using two combination strategies based on the assumption that genetic effects are homogeneous or heterogeneous across studies.

### Combine summary statistics assuming homogeneous effects

We combine the test statistics by summing the scores and their covariance matrices if we assume the genetic effects are homogeneous across all the studies. For example, for *m* = 1, 2 … *M* studies, we sum the score statistics by 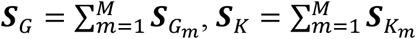 and covariance matrix 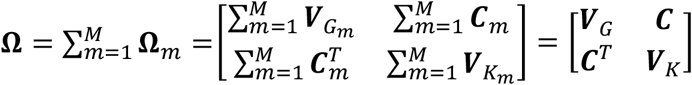. Assuming all participating studies have *q* variants, ***S***_*G*_ is a *q* × 1 vector, ***S***_*K*_ is a *dq* × 1 vector, ***V***_*G*_ is a *q* × *q* matrix, ***V***_*K*_ is a *dq* × *dq* matrix, and ***C*** is a *q* × *dq* matrix. In practice, each variant does not need to be included in all the studies. If a variant is absent in study *m*, we put a placeholder of 0 in place of this variant.

### Combine summary statistics assuming heterogeneous effects

Suppose *B* ethnic groups are studied, some variants can have different allele frequencies depending on the ancestry. Also, lifestyles (e.g., eating habits) and environmental exposures might modify the gene differently, thus their effects on GEI might differ among ethnicities. By correctly accounting for heterogeneity across studies, the power of the meta-analysis should be improved (4). Assuming homogeneous within-ancestry but heterogeneous between-ancestry genetic and GEI effects, we sum the score statistics by 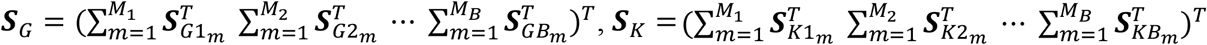 and the covariance matrices by 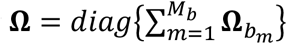 over the studies, where 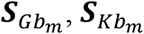, and 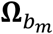 are the genetic score vector, GEI score vector, and covariance matrix of study *m* in ethnic group *b*, respectively. *M*_*b*_ is the number of studies in ethnic group *b* and 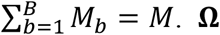 can be partitioned to *diag*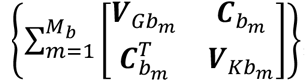, and the variance matrices for genetic effects, GEI effects, and the covariance matrix of genetic and GEI effects are 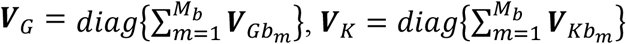, and 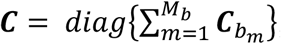, respectively. The lengths are then *Bq* for ***S***_*G*_ and *Bdq* for ***S***_*K*_. ***V***_*G*_, ***V***_*K*_, and ***C*** are block-diagonal matrices with *B* blocks of *q* × *q, dq* × *dq*, and *q* × *dq* sub-matrices respectively for each ancestry.

### GEI tests

In the MAGEE framework (10), we developed two GEI tests: interaction variance component (IV) test and interaction hybrid test using Fisher’s method (IF). The same GEI tests can be reconstructed for the meta-analysis from the combined score vectors and the covariance matrices. IV test assumes the overall GEI effects ***γ*** ∼ *N*(0, *τ****W***_*K*_^2^), for which the test statistic is

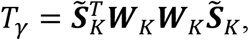

where 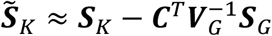 and ***W***_*K*_ is a predefined diagonal weight matrix for interaction. *T*_*γ*_ follows a distribution of 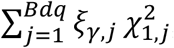, where 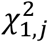 are independent chi-square distributions with 1 degree of freedom (df), and *ξ*_*γ,j*_ are the eigenvalues of ***W***_*K*_**Λ*W***_*K*_, where 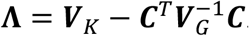.

***W***_*K*_ can be flexibly determined using annotation information or MAFs in the combined studies without having to access the individual-level data (36-38). One of the most popular ways to add weight to variants based on MAF is to use a beta density function with parameters 1 and 25 for the MAF (6). The dimension of ***W***_*K*_ is *Bdq* × *Bdq*, in which *B* = 1 if we combine the summary statistics assuming homogeneous genetic effects.

IF test assumes the overall GEI effects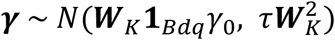, where **1**_*Bdq*_ is a vector of 1’s with length B*dq*. To test for *H*_0_: *γ*_0_ = *τ* = 0, we need two test statistics, 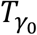 for the burden effects *γ*_0_ and *T*_*τ*_ for the variance component effects *τ*. The burden effects test statistic

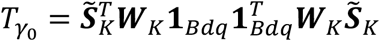

follows the distribution of 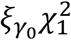 under the null hypothesis *H*_0_: *γ*_0_ = 0, where 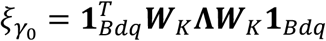. The variance component effects test statistic is

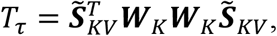

where 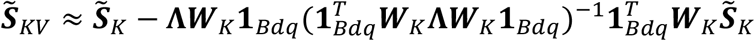. *T*_*τ*_ follows the distribution of 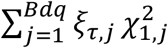 under the null hypothesis *H*_0_: *τ* = 0, where *ξ*_*τ,j*_ are eigenvalues for 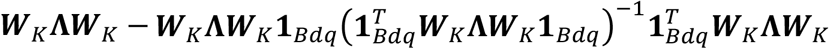. Since 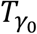 and *T* _*τ*_ are asymptotically independent (proved by Wang *et al*. (10)), a Fisher’s combination (39) can be used to find the *p* value for the IF test: 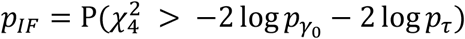, where 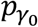 and *p*_*τ*_ are *p* values from the burden-type test *H*_0_: *γ*_0_ = 0 (under the assumption *τ* = 0) and the adjusted variance component test *H*_0_: *τ* = 0, respectively.

### Joint tests

In the joint test, which evaluates both genetic effects and GEI effects, ***β*** and ***γ***, the test statistics for ***β*** must also be reconstructed after summing the summary statistics across all studies. A meta-analysis framework for genetic main effects tests has been developed by SMMAT, which supports both SKAT test and unified burden and SKAT test. Wang *et al*. (10) has shown that those test statistics for the genetic main effects and the GEI test are mutually independent. Assume that the *p* value from the meta-analysis SKAT is *p*_*MV*_ and the *p* value from the meta-analysis IV test is *p*_*IV*_, we can combine them through Fisher’s method as 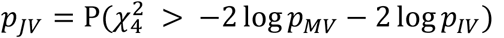 for the joint variance component (JV) test. Similarly, assuming 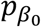 and *p*_*σ*_ are the *p* values from the hybrid genetic main effects meta-analysis, we can combine them with their GEI test counterparts 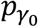 and *p*_*τ*_ through 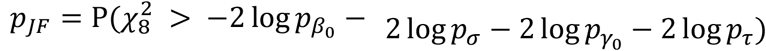, which is the joint hybrid test using Fisher’s method (JF). Alternatively, we can combine the *p* values for genetic effects separately by 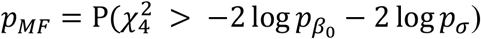, and then combine the main effects and GEI *p* values by 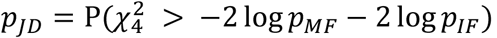, which is the joint hybrid test using double Fisher’s procedures (JD).

### Numerical Simulations

Using related individuals from the same population and different populations, we conducted simulation studies with two purposes: (1) benchmarking *p* values from the meta-analysis to the pooled analysis of individual-level data from MAGEE; and (2) evaluating the performance of each test under the conditions of heterogeneous genetic influence across populations.

We simulated the genotype replicates in two different populations using MS (40) and prepared two scenarios based on the two populations. In scenario 1, we simulated 100,000 individuals from the same population, which included 50,000 parents and 50,000 children in a family relatedness pattern with a kinship matrix of 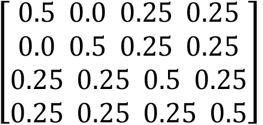. The 100,000 samples were split into two studies of 50,000 individuals (12,500 families) each, and the covariates were the same in the two studies. In scenario 2, we simulated 2 populations with a migration rate of 10. Each population had 50,000 individuals, including 25,000 parents and 25,000 children who were related to each other in the same way as scenario 1. The covariates in these two studies were different. The simulation scenarios and covariates setup for each study are summarized in Table 2. Each scenario had 1,000 genotype replicates of 100 variants in a group. Both qualitative and binary traits were simulated, and the following section describes the details for each simulation.

**Table 2.** Summary of simulation scenarios.

|  | Population 1 | Population 2 | Covariates |
| --- | --- | --- | --- |
| Scenario 1: homogeneous genetic main effects and GEI effects |  |  |  |
| Study 1 | 50,000 | 0 | Age, sex, BMI |
| Study 2 | 50,000 | 0 | Age, sex, BMI |
| Scenario 2: heterogeneous genetic main effects and GEI effects |  |  |  |
| Study 1 | 50,000 | 0 | Age, sex, BMI |
| Study 2 | 0 | 50,000 | Sex, BMI |

### Comparison of *p* values and power

We simulated 10 phenotype replicates per genotype replicate. In each study, the quantitative traits of individual *j* in family *i* were simulated from

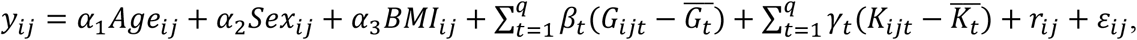

where *Age*_*ij*_ ∼ *N*(50, 5) for parents and *Age*_*ij*_ ∼ *N*(50, 5) for children, *Sex*_*ij*_ ∼ *Bernoulli*(0.5) for both parents and children, BMI for family *i* follows a distribution with heritability (41) of 0.75 (44) ***BMI***_*i*_ ∼ 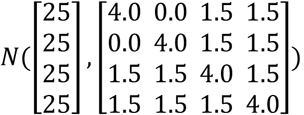, the random effects for family *i* 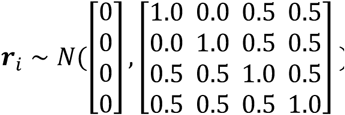, and the random error *ε*_*ij*_ ∼ *N*(0, 1). *G*_*ijt*_ was the *t*-th genetic variant for individual *j* in family *i* and 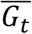 was the mean of the t-th variant, and 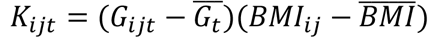 was the GEI term for the *t*-th variant with BMI for individual *j* from family *i* and 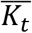 was the mean of the *t*-th interaction term in that study. The binary traits were simulated using a logistic regression model from

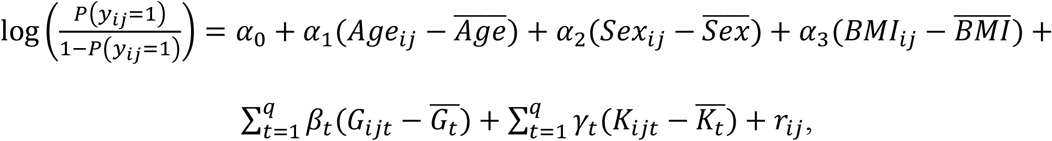

where *Age, Sex, BMI*, and the random effects for family *i* ***r***_*i*_ followed the same distribution as the quantitative traits, and 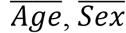, and 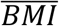 were the population means in each study, *α*_0_ was set to 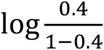 representing a prevalence rate of 0.4. 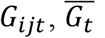 and the calculation of *K*_*ijt*_ and 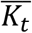 were the same as the quantitative traits.

For scenario 1, the parameters for age, sex, and BMI were *α*_1_ = 0.1, *α*_2_ = 0.2, and *α*_3_ = 0.1, the same for study 1 and study 2. The effect sizes *β*_*t*_ for G and *γ*_*t*_ for GEI were also the same across in both studies. In scenario 2, the parameters for the covariates in study 1 were the same as those for scenario 1, while the parameters in study 2 were *α*_1_ = 0, *α*_2_ = 0.4, and *α*_3_ = 0.12, so that age was excluded from the covariates. In addition, the effect sizes for G and GEI effects were different between the 2 studies in scenario 2. Specifically, the effect sizes of a variant *β*_*t*_ and *γ*_*t*_ were proportional to its MAF as *c*log_10_(*MAF*_*t*_), where c represents a constant. Detailed information of the constants *c* for *β*_*t*_ and *γ*_*t*_ in each simulation can be found in supplemental Table S1 for the *p* value benchmark, supplemental Table S2 for the quantitative trait power comparison, and supplemental Table S3 for the binary trait power comparison.

For the *p* value benchmark, we used different parameters for three different sample sizes. In total, 20% of the variants were randomly selected as causal, which included 10% positive effects and 10% negative effects. For power comparisons, the same parameters were used with different sample sizes, but different parameters were used for GEI and joint tests. We used two approaches to indicate the proportion of causal variants: (1) we randomly chose 20% of causes, among which 10% were positive and 10% were negative; (2) we randomly selected 20% of causes, among which 16% were positive and 4% were negative. The gene-BMI interaction was tested both for binary and quantitative traits. According to Wu *et al*. (6), we used a beta density weight function with parameters 1 and 25 on the MAF, so that rare variants had a higher weight than common variants.

### Application to UK Biobank whole exome sequencing data

We used the UK Biobank (42) whole exome sequencing (WES) data as an example of our proposed meta-analysis. The UK Biobank has released the first tranche of WES data containing 49,959 samples in March 2019. The second tranche of WES data was released in October 2020 with 200,632 samples (including the samples in the first tranche). We divided the dataset in the second tranche by the first and second batches to meta-analyze the two datasets because there may be measurement errors or batch-specific heterogeneities due to differences in quality control procedures. After excluding the subjects with missing sex information, missing age, BMI or WHR, non-white British, and those who have withdrawn from the study, a total of 43,190 subjects were eligible for batch 1 (23,372 women and 19,818 men), and 132,058 subjects were eligible for batch 2 (72,961 women and 59,097 men). The age distribution was mean = 56.95 (*sd* = 7.89) in batch 1 and mean = 56.75 (*sd* = 8.04) in batch 2. The BMI distribution was mean = 27.40 (*sd* = 4.77) in batch 1 and mean = 27.34 (*sd* = 4.70) in batch 2.We tested for gene-by-sex interaction effects on WHR at the gene-level for the two batches. We first fitted a linear mixed model (43) adjusting for sex, age, age^2^ and the top ten ancestry principal components (PCs) for each batch using glmmkin function from the GMMAT package,

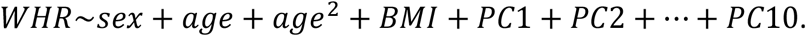

The relatedness matrix was constructed using the kinship coefficients computed by KING software (44) for third-degree and closer relatives, provided by the UK Biobank (see UK Biobank Resource 531).

We used the genotype data generated by the Functionally Equivalent (FE) pipeline (45). The variant groups were defined as protein-coding regions for the WES data and a total of 19,449 protein-coding regions with 17,549,650 variants were available for analysis. MAGEE was used to generate the score statistics and the covariance matrices for each batch. In the next step, we combined and meta-analyzed the scores and covariance matrices for the two batches, assuming homogeneous genetic effects. As IF test was more powerful than the IV test, and JF test was slightly more powerful than JV and JD tests in our power simulations, in the UK Biobank WES data analysis we reported IF and JF test *p* values as the GEI and joint test results, respectively. The meta-analysis excluded variants with a minor allele count (MAC) less than 5, or with a genotype missing rate greater than 5% in the combined dataset, and 18,668 protein-coding regions with 2,328,550 variants passed these filters. We applied a beta density weight function with parameters 1 and 25 on the MAF. In addition, we compared the meta-analysis results to the results of a joint analysis pooling all the individual-level data using MAGEE. For protein-coding regions that reached the Bonferroni-corrected significance level of 0.05/18,668 = 2.7×10^−6^, we also performed conditional analysis by adjusting for variants in the region that had a single variant joint test *p* value less than 5.0×10^−8^, MAF greater than 1% and genotype missing rate less than 5%. Both the genetic main effect and gene-sex interaction terms of significant single variants were included in the conditional analysis.

## Data Availability

The individual-level data that support the findings of this study are available upon application to the UK Biobank (https:/www.ukbiobank.ac.uk/register-apply/).

https:/www.ukbiobank.ac.uk/register-apply/

## Declarations

### Ethics approval and consent to participate

This study was approved by the Institutional Review Board (IRB) of The University of Texas Health Science Center at Houston (UTHealth) with project number HSC-SPH-18-0466. Waiver of informed consent was granted.

### Availability of data and materials

The individual-level data that support the findings of this study are available upon application to the UK Biobank (https://www.ukbiobank.ac.uk/register-apply/).

The method is implemented in R package MAGEE, available at https://github.com/large-scale-gxe-methods/MAGEE.

## Acknowledgements

This research has been conducted using the UK Biobank Resource under Application Number 42646. This work was supported by National Institutes of Health (NIH) grant R01 HL145025.

