## Supplemental Tables and Figures for "Genomic summary statistics and meta-analysis for set-based gene-environment interaction tests in large-scale sequencing studies"

### Supplementary Tables

Table S1 Simulation parameters for constant $c$ in $p$ value benchmarking

| | | **Quantitative trait** | | **Binary trait** | |
|---|---|---|---|---|---|
| | | $\beta_t$ | $\gamma_t$ | $\beta_t$ | $\gamma_t$ |
| Scenario 1: homogeneous genetic main effects and GEI effects | | | | | |
| Study 1 & Study 2 | | | | | |
| Sample size | 20,000 | 0.012 | 0.013 | 0.033 | 0.036 |
| | 50,000 | 0.009 | 0.009 | 0.018 | 0.02 |
| | 100,000 | 0.007 | 0.0072 | 0.009 | 0.012 |
| Scenario 2: heterogeneous genetic main effects and GEI effects | | | | | |
| Study 1 | | | | | |
| Sample size | 20,000 | 0.024 | 0.03 | 0.06 | 0.066 |
| | 50,000 | 0.018 | 0.019 | 0.04 | 0.045 |
| | 100,000 | 0.008 | 0.0095 | 0.018 | 0.02 |
| Study 2 | | | | | |
| Sample size | 20,000 | 0.008 | 0.01 | 0.02 | 0.022 |
| | 50,000 | 0.006 | 0.0062 | 0.012 | 0.015 |
| | 100,000 | 0.0025 | 0.0032 | 0.006 | 0.007 |

Table S2 Simulation parameters for constant $c$ in power comparison on continuous traits

| | GEI test | | Joint test | |
|---|---|---|---|---|
| | $\beta_t$ | $\gamma_t$ | $\beta_t$ | $\gamma_t$ |
| Scenario 1: homogeneous genetic main effects and GEI effects; +/0/-: 10%/80%/10% of causal variants | | | | |
| Study 1 & Study 2 | 0.04 | 0.028 | 0.052 | 0.022 |
| Scenario 1: homogeneous genetic main effects and GEI effects; +/0/-: 16%/80%/4% of causal variants | | | | |
| Study 1 & Study 2 | 0.038 | 0.028 | 0.05 | 0.022 |
| Scenario 2: heterogeneous genetic main effects and GEI effects; +/0/-: 10%/80%/10% of causal variants | | | | |
| Study 1 | 0.08 | 0.064 | 0.12 | 0.048 |
| Study 2 | 0.03 | 0.02 | 0.04 | 0.018 |
| Scenario 2: heterogeneous genetic main effects and GEI effects; +/0/-: 16%/80%/4% of causal variants | | | | |
| Study 1 | 0.06 | 0.054 | 0.1 | 0.04 |
| Study 2 | 0.02 | 0.018 | 0.035 | 0.015 |

Table S3 Simulation parameters for constant $c$ in power comparison on binary traits

| | GEI test | | Joint test | |
|---|---|---|---|---|
| | $\beta_t$ | $\gamma_t$ | $\beta_t$ | $\gamma_t$ |
| Scenario 1: homogeneous genetic main effects and GEI effects; +/0/-: 10%/80%/10% of causal variants | | | | |
| Study 1 & Study 2 | 0.1 | 0.1 | 0.135 | 0.08 |
| Scenario 1: homogeneous genetic main effects and GEI effects; +/0/-: 16%/80%/4% of causal variants | | | | |
| Study 1 & Study 2 | 0.1 | 0.1 | 0.14 | 0.085 |
| Scenario 2: heterogeneous genetic main effects and GEI effects; +/0/-: 10%/80%/10% of causal variants | | | | |
| Study 1 | 0.36 | 0.32 | 0.4 | 0.25 |
| Study 2 | 0.12 | 0.1 | 0.15 | 0.08 |
| Scenario 2: heterogeneous genetic main effects and GEI effects; +/0/-: 16%/80%/4% of causal variants | | | | |
| Study 1 | 0.35 | 0.31 | 0.39 | 0.24 |
| Study 2 | 0.11 | 0.1 | 0.13 | 0.08 |

### Supplementary Figures

Figure S1. GEI test *p* value benchmark for meta-analysis assuming homogeneous genetic effects with MAGEE in 20,000, 50,000, and 100,000 unrelated samples. (A) Scenario 1 (homogeneous scenario) in binary traits. (B) Scenario 2 (heterogeneous scenario) in binary traits.

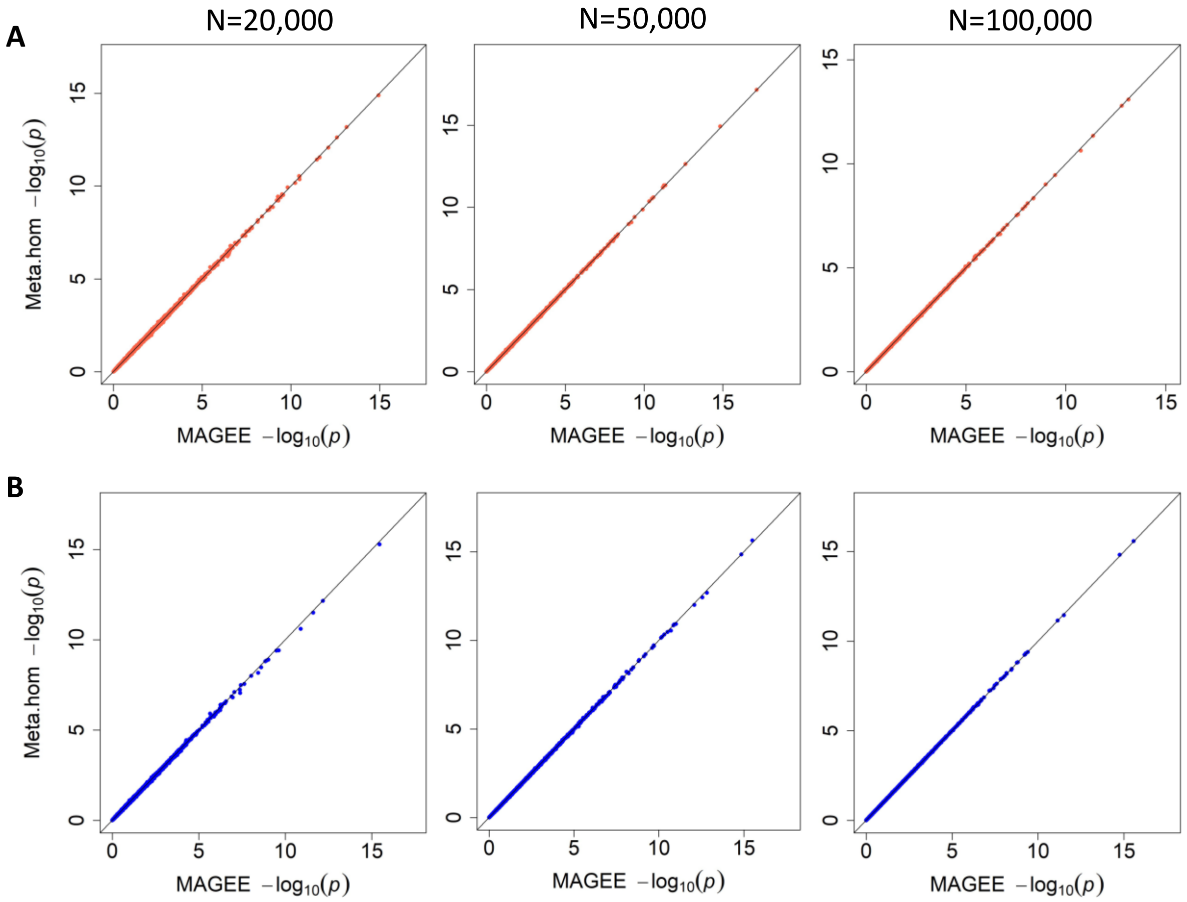

Figure S2. Joint test *p* value benchmark for meta-analysis assuming homogeneous genetic effects with MAGEE in 20,000, 50,000, and 100,000 unrelated samples. (A) Scenario 1 (homogeneous scenario) in binary traits. (B) Scenario 2 (heterogeneous scenario) in binary traits.

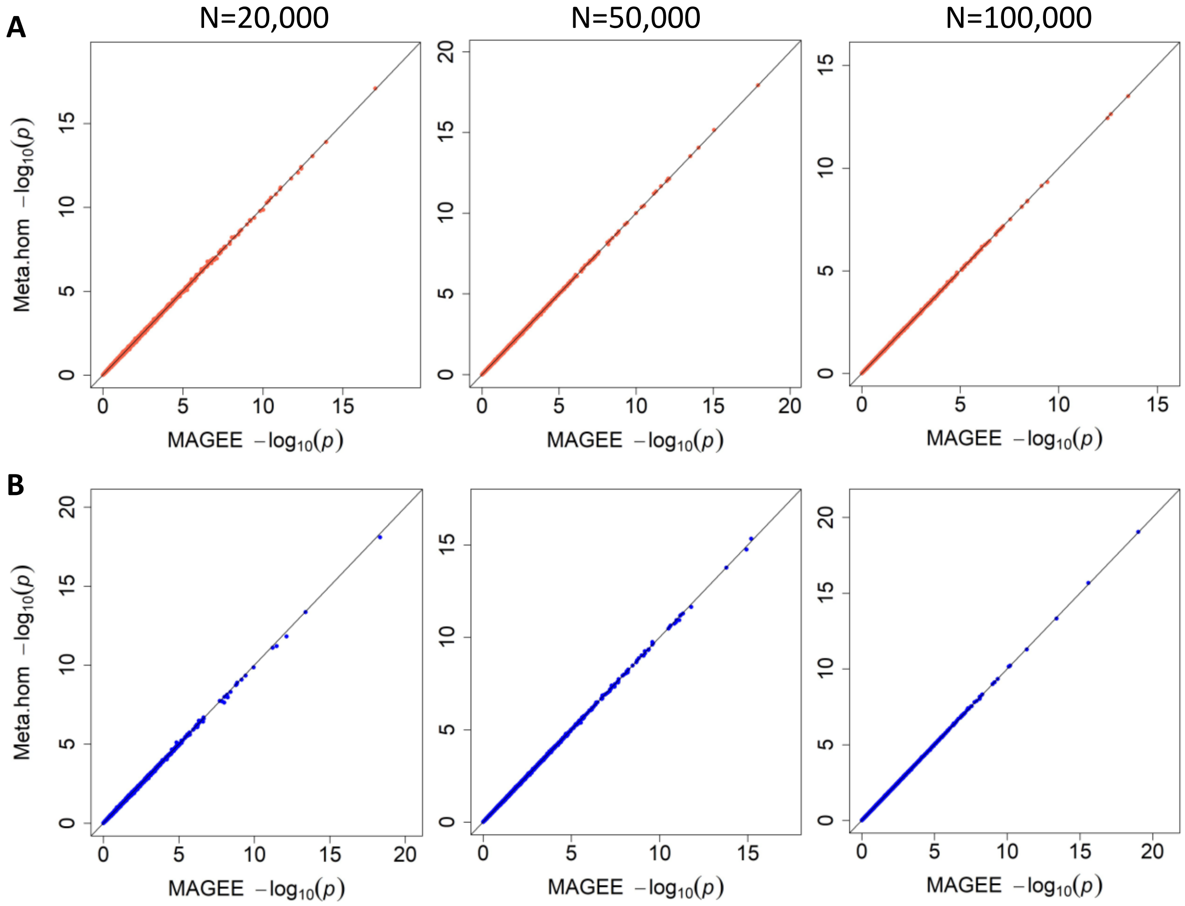

Figure S3. Empirical power of meta-analysis tests for scenario 1 (homogeneous scenario) on binary traits in 20,000, 50,000, and 100,000 related samples. (A) GEI tests with 80% null variants, 10% causal variants with positive effects, and 10% causal variants with negative effects. (B) Joint tests with 80% null variants, 10% causal variants with positive effects, and 10% causal variants with negative effects. (C) GEI tests with 80% null variants, 16% causal variants with positive effects, and 4% causal variants with negative effects. (D) Joint tests with 80% null variants, 16% causal variants with positive effects, and 4% causal variants with negative effects.

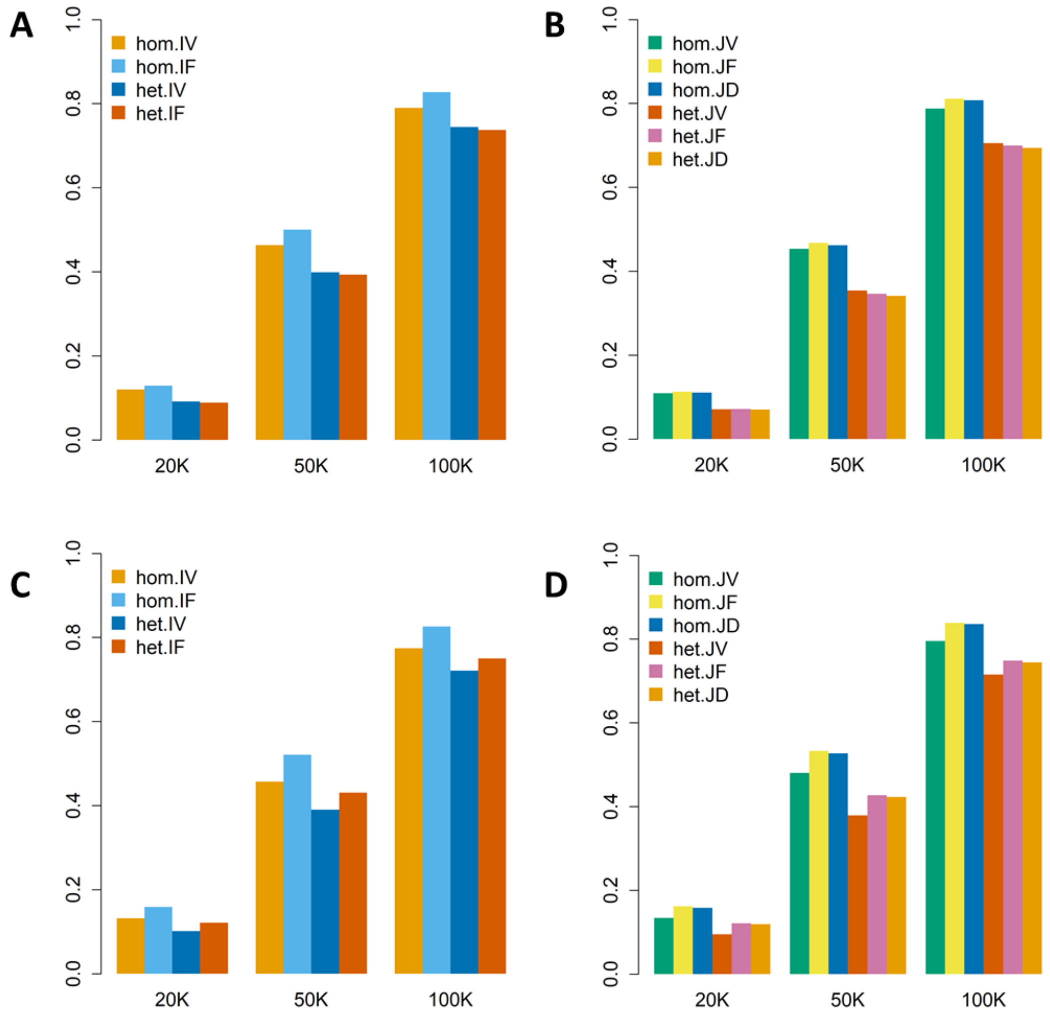

Figure S4. Empirical power of meta-analysis tests for scenario 2 (heterogeneous scenario) on binary traits in 20,000, 50,000, and 100,000 related samples. (A) GEI tests with 80% null variants, 10% causal variants with positive effects, and 10% causal variants with negative effects. (B) Joint tests with 80% null variants, 10% causal variants with positive effects, and 10% causal variants with negative effects. (C) GEI tests with 80% null variants, 16% causal variants with positive effects, and 4% causal variants with negative effects. (D) Joint tests with 80% null variants, 16% causal variants with positive effects, and 4% causal variants with negative effects.

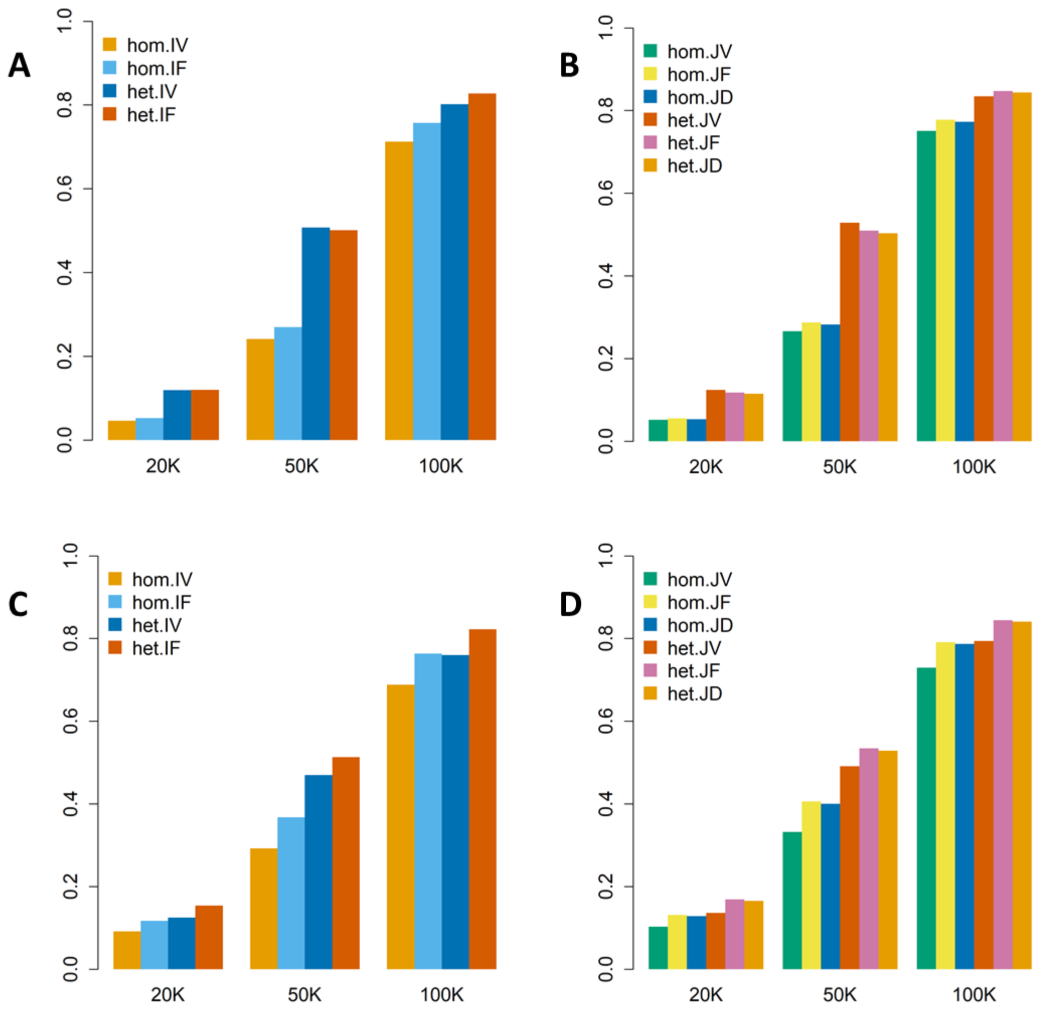

Figure S5. Scatter plots comparing MAGEE pooled analysis results using individual-level data, and meta-analysis results from gene-sex interaction analysis on WHR using UK Biobank WES data. (A) GEI test. (B) Joint test. Spearman’s rank correlation coefficient was shown on the top left of each panel.

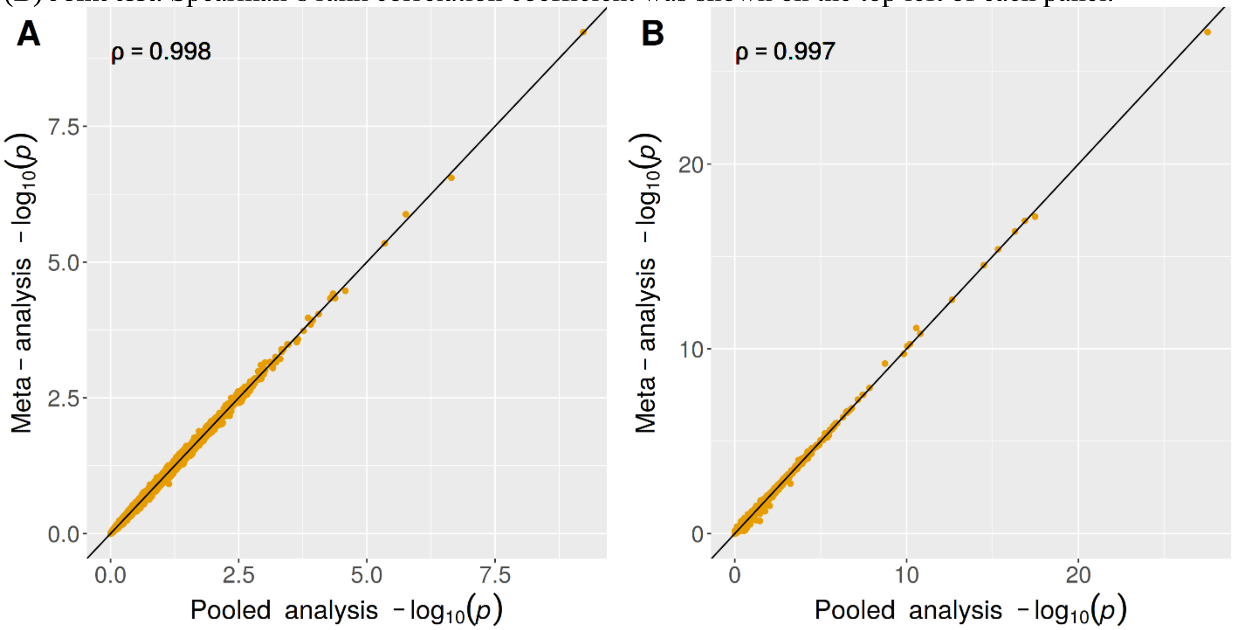
